# Effectiveness of technological interventions to improve upper limb motor function in people with stroke in low- and middle-income countries: Protocol for a systematic review and meta-analysis

**DOI:** 10.1101/2020.11.10.20209197

**Authors:** Meiling Milagros Carbajal-Galarza, Nathaly Olga Chinchihualpa-Paredes, Sergio Alejandro Abanto-Perez, María Lazo-Porras

**Affiliations:** School of Medicine. Universidad Peruana Cayetano Heredia. Lima-Perú; CRONICAS Centre of Excellence in Chronic Disease. Universidad Peruana Cayetano Heredia. Lima-Perú; CONEVID. Unidad de Conocimiento y Evidencia. Facultad de Medicina Alberto Hurtado. Universidad Peruana Cayetano Heredia; Division of Tropical and Humanitarian Medicine, Geneva University Hospitals & University of Geneva, Switzerland

**Keywords:** stroke, technology, upper extremity, low-and middle- income countries

## Abstract

**Introduction:** Stroke is one of the main causes of disability in low- and middle-income countries (LMIC), frequently presenting with upper extremity paresis and causing major functional dependence. It requires high dose and intense rehabilitation which implies high economic costs, consequently limiting this therapy in LMIC. There are multiple technological interventions that facilitate rehabilitation either in intensity, adherence and motor evaluation; or enable access to rehabilitation such as robots, games or virtual reality, sensors, electronic devices and tele-rehabilitation. Their efficacy has been mainly evaluated in high-income countries, hence the importance of conducting a systematic review in LMIC settings.

**Objectives:** To measure the efficacy of technological interventions vs. classical physical rehabilitation in the upper extremity motor function in people who had suffered a first or recurrent episode of stroke in LMIC.

**Methods and analysis:** This protocol is consistent with the methodology recommended by the PRISMA-P and the Cochrane handbook for systematic reviews of interventions. We propose to do a systematic review and meta-analysis. In order to do so, we will perform an electronic search in PubMed, Global Index Medicus and Physiotherapy Evidence Database. No date range parameters will be used. Randomized controlled trials (RCT) published in English, Spanish, French and Portuguese, with the primary outcome focusing on upper limb motor function, will be included. Two reviewers will screen all retrieved titles, abstracts and full texts, perform the evaluation of the risk of bias and extract all data independently. The risk of bias of the included RCT will be evaluated by the Cochrane Collaboration’s tool. A qualitative synthesis will be provided in text and tables, to summarize the main results of the selected publications.The heterogeneity between studies will be assessed through the I^2^ statistic. If there is sufficient homogeneity across outcomes, a meta-analysis will be considered. The outcomes to be evaluated will be motor functionality of the upper extremity, performance for activities of daily living and quality of life, through measurement scales.

**Conclusions:** This systematic review will provide evidence regarding the efficacy of multiple technological interventions to improve motor function of upper extremity in individuals with stroke in LMIC. Based on this analysis, we will be able to assess whether these interventions are also effective and feasible in the recovery of functionality after stroke in low- and middle-income countries, and thus offer recommendations in these areas.

## INTRODUCTION

Stroke is one of the worldwide leading causes of disability in adults.Low- and middle-income countries (LMIC) have the highest burden of stroke in the world, plus higher death rates and disability-adjusted life years than high-income countries (HIC). Besides, while stroke incidence rates are declining in HIC, the opposite is occurring in LMIC.^1^ Among the factors contributing to this increase in LMIC is rapid urbanization, lengthening of life expectancy, undiagnosed hypertension and increased tobacco use.^1,2^ However, most of the research on stroke rehabilitation, including research that informs guidelines for good clinical practices, has been conducted in HIC.^2^

One of the most frequent consequences of stroke is the movement and coordination impairment of the upper extremity with a prevalence of 55% to 75% in patients suffering upper extremity motor deficit between 3 and 6 months after the stroke ^3^, of whom only a small part recover completely, resulting in a majority with persistent disability and restrictions in their activities of daily living.^4,5^ The upper limb limitation results in functional dependence, loss of important occupations and may even lead to institutionalization.^6^ Likewise, it has been shown that the recovery of upper extremities is harder to achieve compared to lower extremities, mainly due to the complexity of the functions performed by the first.^5^ Particularly hand rehabilitation, due to its complex structure, specialized functions and intricate neural base. ^7^ Therefore, the importance of multidisciplinary rehabilitation after stroke and research for new therapies and strategies to regain motor skills and improve functional outcomes in upper extremities.Neuroplasticity is the basic mechanism underlying rehabilitation, its mechanisms involved put into operation pre-existing synaptic connections, modify synaptic connections determined for neuronal activity, and develop new connections or assume the lost function by uninjured adjacent cortical areas.^5^ Other principles of stroke rehabilitation include goal setting, high intensity practice, and training for specific tasks. Therefore, intensive training in high doses and repetitive practice of specific tasks is essential in recovery after stroke.^8,9^

The main objective of rehabilitation technologies is to maintain or improve the functioning and independence of an individual, in addition to facilitating participation and improving general well-being.^10^ In the last decade, the use of technological devices in the rehabilitation of patients affected by stroke has increased, which are developed to improve stroke results, due to the increase in intensity, repetitions, specificity and feedback during rehabilitation.^10,11^ There is a wide range of technologies with applications in rehabilitation. Iosa conducts a review describing the “Seven capital devices for the future of Stroke rehabilitation”: robots, brain computer interfaces, neuroprostheses, non-invasive brain stimulators, wearable devices, virtual reality and tablet-pc.^11^ Likewise, other technologies such as assistive devices, video games, use of mobile health platforms and telehealth are reported.^12^

Robotic rehabilitation therapy offers high-intensity and high-dose training.^13^ It is classified into end-effector-based and exoskeleton-type robots. There are multiple brands such as MIT - Manus, MIME, ARM, GENTLE, NeReBot, ARMin, among others.^8^ A 2012 Cochrane review found that robotic therapy of the upper limb provides benefits with respect to activities of daily living and arm function, but not on arm muscle strength.^14^ Another systematic review shows, that robotic therapy presents uncertain utility with minimal differences when compared with conventional therapy matched in intensity and dose, however, when combining extra sessions of robotic therapy with conventional therapy, these are more beneficial in the motor recovery of the paretic shoulder and elbow than conventional therapy alone.^13^ The 2016 American Heart Association Adult Stroke Recovery and Rehabilitation Guide suggests a class IIa level of evidence for robotic therapy, which offers more intensive practice to people with moderate to severe limb paresis.^6^ However, the majority of studies conducted regarding robotics have been carried out in developed countries and the efficacy and viability of these in lower income countries has not been proven.

Virtual reality offers increased training intensity and repetitions, as well as reducing the therapist’s time.^10^ Likewise, video games and virtual reality have the potential to increase participant participation by making repetitive exercises more engaging and motivating.^15^ A Cochrane review finds that the use of virtual reality and interactive video games was not more beneficial than conventional therapy approaches to improve upper limb function. However, it may be beneficial in improving upper limb function and activities of daily living when used as an adjunct to regular care.^16^ This review was conducted mostly in high-income countries, and no differentiation was made in the results in those low- and middle-income countries considered.

Telerehabilitation uses information and telecommunications technologies to help patients receive medical services from remote healthcare providers, through video conferencing that allows therapists to observe the movement of patients when performing rehabilitation tasks. This technology is especially useful in stroke patients who have difficulties with transportation, depend on caregivers or lack rehabilitation services in their geographic areas; which are more frequent situations in low-income countries.^15^ A meta-analysis provided limited and moderate evidence on telerehabilitation showing that it has the same effects as conventional rehabilitation in improving the skills of activities of daily living and motor function of stroke survivors.^17^ Because of its special utility in low-income countries, it is important to determine its efficacy in this setting.

There are other technologies that have not been evaluated for their effectiveness in low- and middle-income countries either. An example are brain-computer interfaces, which are a family of various devices that aim to translate brain activity into external device control signals that can replace, complement, or enhance normal neural output.^11,18^ In the case of EEG-based brain-computer interfaces, the user’s movement intent is decoded in real time from the brain’s ongoing electrical activity, then the movement intent detection triggers tangible sensory feedback for the user which can be delivered in an abstract way such as through the movement of a cursor on the screen or feedback incorporated with other technologies such as virtual reality, robots, among others.^18^ Neuroprosthesis or Functional electrical stimulation are electronic devices that involve electrical stimulation of motor neurons, causing contraction of muscle groups to create or augment momentum around a joint.^11,19^ Another technology in rehabilitation is non-invasive brain stimulation, which consists of 2 techniques: repetitive transcranial magnetic stimulation and direct current transcranial stimulation^11^ that use electric currents or magnetic fields directed towards the frontal cortex to modulate motor cortical function.^20^ There are several encouraging studies that have demonstrated functional improvement, however, its efficacy and cost-effectiveness, either as home therapy or hospital rehabilitation, have not been evaluated in low- and middle-income countries.

### Justification

Stroke is one of the main causes of disability in LMIC, with upper extremity paresis as a very frequent disability, which generates great functional dependence, and is the most complicated to recover its function. Therefore, it requires intensive rehabilitation, at a high dose and feedback. However, this implies large economic costs not only in the type of rehabilitation used, but also in the costs inherent to the care provided such as transportation of the patient or the therapy itself (salary of the therapist, place of rehabilitation, etc.), which often limits the duration and frequency of therapies.^21^ There are various technologies in rehabilitation that seek both to improve the functionality of the upper extremity and to reduce barriers in rehabilitation such as robots, which increase the intensity and dose of rehabilitation; video games and virtual reality, which encourage adherence; electronic devices such as sensors, which allow distant motor assessment; tele-rehabilitation that facilitates access to rehabilitation; brain-computer interfaces, which promote feedback; and non-invasive brain stimulation, which seeks to modulate cortical motor function.^6,22^ However, the efficacy of these has been evaluated mostly in high-income countries, and there are no systematic reviews on the efficacy of these technological interventions in upper extremity rehabilitation in LMIC.

### Objectives

#### Primary objective

- To evaluate the efficacy of technological interventions vs classical physical rehabilitation with respect to upper extremity motor function in people who have suffered a first or recurrent episode of stroke in low- and middle- income countries.

#### Secondary objectives

- To assess the efficacy of technological interventions in relation to independence in the execution of activities of daily living of people who have suffered a first or recurrent episode of stroke in low- and middle- income countries.
- To assess the efficacy of technological interventions in the quality of life of people who have suffered a first or recurrent episode of stroke in low- and middle- income countries.

## METHODS AND ANALYSES

### Study design

This review protocol was written and reported following the Preferred Reporting Items for Systematic Reviews and Meta-Analysis Protocols (PRISMA-P).^23^ The results of this systematic review will be published following the PRISMA statement.^24^

### Eligibility criteria

#### Types of study

We will include all randomized controlled trials (RCT) published in English, Spanish, French and Portuguese that investigated the efficacy of different types of technological interventions to improve upper extremity motor function in individuals who have suffered from stroke in LMIC.

It should be noted that studies that have only evaluated the efficacy of technological interventions in relation to independence in the execution of activities of daily living (ADL), and in relation to the quality of life of people who have suffered an episode of cerebrovascular accident, will also be included in order to assess secondary objectives. In the case of multicenter studies, they will be included only if the results are disaggregated and differentiated by country. This to select only the information from LMIC, excluding HIC. In addition, these last studies will also be considered if the published results are not disaggregated by country, but can be obtained through contact with the authors.

#### Participants

We will include all RCTs which have recruited adult patients (≥ 18 years of age) with a first or recurrent episode of stroke living in LMIC.

Stroke is defined as ‘a clinical syndrome consisting of rapidly developing clinical signs of focal (or global in case of coma) disturbance of cerebral function lasting more than 24 hours or leading to death with no apparent cause other than a vascular origin’ by WHO. ^25^ This term involves cerebrovascular accidents of ischemic and hemorrhagic origin.

The diagnosis of stroke should have been confirmed by CT or MRI in all these patients. Furthermore, the participants in all trials should have been assessed with at least one of the selected scales for upper extremity motor dysfunction and upper extremity functionality, as well as scales that evaluate the independence for activities of daily living and the quality of life in patients who have suffered a stroke event. These scales are more specifically detailed in the outcomes section. Patients with subarachnoid hemorrhage or subdural hematoma will be excluded. Likewise, studies with participants with transient ischemic attack will be excluded since all neurological symptoms would disappear.

#### Types of intervention

We will select all trials that evaluated interventions where at least one of the treatment goals aims at recovery of upper extremity function after a stroke.

We will address individual interventions in rehabilitation using technological devices ^6-22^ for the upper extremities such as robots, video games and virtual reality, telerehabilitation, wearable devices, neuroprosthetics or functional electrical stimulation, repetitive transcranial magnetic stimulation, transcranial direct current stimulation and brain-computer interfaces.

Interventions could have been performed individually or in groups, in the hospital or at home, supervised by therapists or self-training. No limits will be imposed on the timing, frequency, and duration of the interventions.

#### Comparison

These interventions should be compared with a control intervention like no treatment, standard care, placebo treatment or conventional rehabilitation.

#### Types of outcome measures

Several scales were selected and studied in the domains of disability ^26-28^ and quality of life ^29-30^, taking into consideration the International Classification of Functioning, Disability and Health.^31-33^

The primary results of this systematic review will focus on changes in patients’ upper extremity functionality following the intervention. The assessments of arm functioning are divided into two types:

□ Motor impairment of the arm (measures of sensorimotor disability of the upper limb, muscle strength, muscle tone): Fugl-Meyer Assessment of Sensorimotor Recovery after Stroke (upper limb section) ^34^, Motricity Index ^35^, Chedoke-McMaster Stroke Assessment ^36^.
□ Functional movement of the upper limb that assess activity limitations (such as measures of active movement, coordination, dexterity, manipulation, grasp/grip) which are:
  □ Arm Function: Action Research Arm Test ^37^, Wolf Motor Function Test ^38^, Box and Block Test ^39^, Chedoke Arm and Hand Activity Inventory.^40^
  □ Hand Function: Purdue Peg Test ^41^, Nine Hole Peg Test ^42^, Motor Assessment Scale - hand movement or advanced hand movement scores ^43^.

Secondary outcome measures will include the following scales:

□ Independence in Activities of Daily Living (ADLs) (including feeding, dressing, bathing, simple mobility and transfers): Barthel Index ^44^, Rankin Scale ^45^ and Functional Independence Measure ^46,47^.
□ Quality of life: Medical Outcomes Study Short-Form 36-item survey (SF-36) ^48^, Stroke Specific Quality of Life Scale (SS-QOL) ^49^, Stroke Impact Scale version 3.0 (SIS 3.0) ^50^.

#### Time frame

We will not include a time frame or limitation with respect to time for the development of this systematic review.

### Exclusion criteria

- The following types of publications will be excluded: uncontrolled, non-randomized trials, pilots, case reports, case series, letter to the editor, editorial, observational and epidemiological studies, narrative review and systematic reviews.
- We will exclude documents in languages other than Spanish, English, Portuguese and French, as well as studies that are not accessible in full text despite having been requested from the author.
- We will exclude studies with participants under 18 years of age or that refer to other entities of brain injury such as subarachnoid hemorrhage, subdural hematoma or transient ischemic attack.
- We will exclude studies that are conducted in HIC. Studies conducted in both high-income and low-income countries that do not present country-disaggregated results, despite a request to the author, will be excluded.
- We will exclude studies with only standard or conventional intervention without the use of technological devices.

### Information sources

Electronic searches will be performed for potentially eligible RCTs in the PubMed, Global Index Medicus: African Index Medicus - AIM (AFRO/WHO), Latin American and Caribbean Scientific and Technical Literature - LILACS (AMRO-PAHO/WHO), Index Medicus for the Eastern Mediterranean Region - IMEMR (EMRO/WHO), Index Medicus for the South East Asian Region - IMSEAR (SEARO/WHO), Index Medicus for the Western Pacific Region - WPRIM (WPRO/WHO), and Physiotherapy Evidence databases with restriction in articles with full texts in English, Spanish, French and Portuguese.

We will use a combination of the following keywords in English: stroke, paresis, technology, rehabilitation, stroke rehabilitation, virtual reality, robotics, telerehabilitation, video games, developing countries, low- and middle- income countries, randomized controlled trial, controlled clinical trial.

### Search strategy

A different search strategy will be designed for each database. The complete preliminary search strategy of these databases can be found in Appendix 1 (Supplementary material).

### Study records

#### Data management

The reference management software, Rayyan QCRI, will be used to help upload, store and select the literature results. For each database, a separate library group will be created to keep all original search results. All separate library group copies will then merge into a new library group and duplicate checking will be carried out.

#### Selection process

The selection process will be carried out by two independent reviewers according to the title and abstract of each study, in order to find relevant ones. This will be done considering the inclusion and exclusion criteria previously detailed. Subsequently, once more, two independent reviewers will look into the full text of previously selected articles to confirm the final selection. Any conflict will be solved by agreement or consulting a third author.All publication exclusions will be justified. The PRISMA flow of information through the different phases of a systematic review will be filled in, to record the whole screening process in detail.

#### Data collection process

Two independent reviewers will carry out the data extraction through Google Forms where the variables of interest will be included, then this information will be exported to Microsoft Excel 2019 sheets. The extracted data will include general study information, characteristics of participants, characteristics of interventions, and outcome measures, which are detailed in the data items section. If necessary, the corresponding authors of the selected publications will be contacted for missing data and further information. Disagreements between the two reviewers will be solved by a third reviewer to reach a consensus.

### Data items

The extracted data will include:

- Study details: First author, corresponsal author, article title, multicountries characteristics, country/countries, publication year, language
- Participant characteristics: Sample size (number of participants in each group/shot/arm), inclusion and exclusion criteria, age range, gender, randomization process, stroke details (hemisphere, time after stroke, type of stroke)
- Details of interventions: type of intervention, duration, frequency, supervision, implementer, arm segment, control or comparison group
- Outcome measures: observation time, analysis of upper extremity motor functioning (scale used, pre- and post-treatment results in intervention and control group), changes in independence of activities of daily living (scale used, pre- and post-treatment results in intervention and control group), changes in quality of life (scale used, pre- and post-treatment results in intervention and control group), desertion (overall and separated by intervention and control group), duration of follow-up

If an article includes high-income country participants in the sample or if any required data is missing or unclear, we will try to contact the corresponding author. If we do not have a response from the corresponding author and the article does not report the results of our relevant population separately, we will exclude the study.

If data from a study was reported in more than one article, we will only select the article with the most complete data for our research purposes.

### Outcomes and prioritization

The main or initial goal of many upper limb rehabilitation interventions is to improve upper limb functionality, while improving functional movement and reducing impairment. Therefore, it will be considered our primary outcome of interest. On the other hand, it is questionable how significant these aspects are for individual patients, being an important objective to achieve independence with daily life activities and quality of life. Hence, both will be considered secondary outcomes of interest for analysis.

For each outcome of interest (primary and secondary) we propose to identify and list all common and specific measurement instruments or scales that can be included. If a study presents a secondary outcome of interest, it will be considered for such analysis. If we identify a study that reports on more than one measurement instrument or scale addressing the same outcome, we will use the scale listed first. However, if these scales are not available, we will accept other scales that measure arm and hand functioning, independence in activities of daily living, and quality of life.

### Risk of bias in individual studies

We will assess the risk of bias using different Cochrane Collaboration tools. In this case, risk of bias (ROB) will be used for randomized clinical trials, which assesses the following domains: Biases in the randomization process, due to deviations from the planned interventions, due to incomplete results data, in the measurement of results and in the selective reporting of results. It will classify them as low risk of bias, some concerns, and high risk of bias.^51^

These risks of bias judgments will be made by two independent reviewers and disagreements will be resolved first by discussion and then by referring to a third review author as a referee when necessary.

### Strategy for data synthesis

We will provide a qualitative synthesis, in text and table, to summarize the main results of the selected publications. A narrative synthesis will be included to demonstrate the findings, structured around the general characteristics of the study, characteristics of the target population, details of the intervention and types of results. We will check the heterogeneity of the included studies by performing the I^2^ statistical test (high levels of heterogeneity: I^2^ ≥ 75%). For studies that have sufficient data and are homogeneous in terms of interventions and outcomes, we will synthesize the results in meta-analyses using Review Manager software, Comprehensive Meta-analysis, StatsDirect, or MetaDisc software. The random effects model will be used for the combination of results, taking into account heterogeneity. Finally, a forest plot will be carried out with the result of the meta-analysis and, if possible, we will carry out a sensitivity analysis using the funnel plot and the Egger test. In case of substantial heterogeneity, only qualitative synthesis will be performed.^52^

Subgroup analysis will be performed if sufficient data is available. These analyses will involve differences between the income level of countries (low- and middle-income countries / middle-income countries / low-income countries) and details of the intervention (type, duration and execution of the intervention).

### Quality of evidence

If possible, the quality of evidence will be assessed using the GRADE approach for each outcome using the GRADEpro tool. This system considers the risk of bias within the study, the consistency of the effect, indirect evidence, precision of the effect estimates, and publication bias. The overall quality of the evidence will be rated at four levels: high, moderate, low, and very low. ^53^

## Data Availability

For supplementary material online of this manuscript, the next link is provided

https://drive.google.com/drive/folders/1t6oMhfP9kh8YG7HPIftN5lztM891KZVh?usp=sharing

## ETHICS AND DISSEMINATION

This systematic review does not require ethical approval or informed consent. The results of this review will be disseminated through peer-reviewed publications.

## ACKNOWLEDGEMENT

We want to acknowledge Ms. Patricia Busta Flores for the help in the revision of this manuscript in English.

## Appendix

### Appendix 1. Preliminary search strategies

#### A. Pubmed

##### Descriptor 1: Stroke

458199 results

###### Mesh terms

(((“Stroke”[Mesh] OR “Stroke, Lacunar”[Mesh] OR “Infarction, Posterior Cerebral Artery”[Mesh] OR “Brain Stem Infarctions”[Mesh] OR “Infarction, Middle Cerebral Artery”[Mesh] OR “Infarction, Anterior Cerebral Artery”[Mesh] OR “Cerebral Hemorrhage”[Mesh] OR “Brain Ischemia”[Mesh]) OR “Paresis”[Mesh]) OR “Hemiplegia” [Mesh])

###### Free terms

“Stroke” [Majr] OR “stroke*” OR “strokes” OR “Brain Infarction” OR “brain infarctions” OR “cerebrovascular disorder” OR “cerebrovascular disorders” OR “cerebrovascular accident” OR “cerebrovascular accidents” OR “cva” OR “cvas” OR “cerebral infarction” OR “cerebral infarctions” OR “Intracranial Embolism and Thrombosis” OR “cerebrovascular disease” OR “cerebrovascular diseases” OR “cerebral haemorrhage” OR “cerebral hemorrhage” OR “brain ischemia” OR “brain ischaemia” OR “cerebral ischemia” OR “cerebral ischaemia” OR “brain vascular accident” OR “cerebrovascular Stroke” OR “Cerebral Stroke” OR “Acute Stroke” OR “Acute Strokes” OR “basal ganglia cerebrovascular disease” OR “intracranial arterial diseases” OR “intracranial embolism and thrombosis” OR “intracranial hemorrhages” OR “poststroke” OR “apoplex*” OR “cerebral vasc*” OR “brain vasc*” OR “Paresis” OR “Pareses” OR “Muscular Paresis” OR “Muscular Pareses” OR “Muscle Paresis” OR “Muscle Pareses” OR “Hemiparesis” OR “Hemipareses” OR “hemiplegia” OR “hemipleg*”OR “hemipar*” OR “paretic*”

###### Mesh terms + Free terms

((((“Stroke”[Mesh] OR “Stroke, Lacunar”[Mesh] OR “Infarction, Posterior Cerebral Artery”[Mesh] OR “Brain Stem Infarctions”[Mesh] OR “Infarction, Middle Cerebral Artery”[Mesh] OR “Infarction, Anterior Cerebral Artery”[Mesh] OR “Cerebral Hemorrhage”[Mesh] OR “Brain Ischemia”[Mesh]) OR “Paresis”[Mesh]) OR “Hemiplegia” [Mesh]) OR (“Stroke” [Majr] OR “stroke*” OR “strokes” OR “Brain Infarction” OR “brain infarctions” OR “cerebrovascular disorder” OR “cerebrovascular disorders” OR “cerebrovascular accident” OR “cerebrovascular accidents” OR “cva” OR “cvas” OR “cerebral infarction” OR “cerebral infarctions” OR “Intracranial Embolism and Thrombosis” OR “cerebrovascular disease” OR “cerebrovascular diseases” OR “cerebral haemorrhage” OR “cerebral hemorrhage” OR “brain ischemia” OR “brain ischaemia” OR “cerebral ischemia” OR “cerebral ischaemia” OR “brain vascular accident” OR “cerebrovascular Stroke” OR “Cerebral Stroke” OR “Acute Stroke” OR “Acute Strokes” OR “basal ganglia cerebrovascular disease” OR “intracranial arterial diseases” OR “intracranial embolism and thrombosis” OR “intracranial hemorrhages” OR “poststroke” OR “apoplex*” OR “cerebral vasc*” OR “brain vasc*” OR “Paresis” OR “Pareses” OR “Muscular Paresis” OR “Muscular Pareses” OR “Muscle Paresis” OR “Muscle Pareses” OR “Hemiparesis” OR “Hemipareses” OR “hemiplegia” OR “hemipleg*”OR “hemipar*” OR “paretic*”)

##### Descriptor 2: Rehabilitation

11,800,232 results

###### Mesh terms

((“Rehabilitation”[Mesh] OR “Physical and Rehabilitation Medicine”[Mesh] OR “Stroke Rehabilitation”[Mesh] OR “Neurological Rehabilitation”[Mesh] OR “Orthopedic Procedures”[Mesh] OR “Telerehabilitation”[Mesh] OR “Exercise Therapy”[Mesh] OR “Treatment Outcome”[Mesh] OR “Rehabilitation Centers”[Mesh] OR “rehabilitation” [Subheading] OR “Activities of Daily Living”[Mesh] OR “Recreation Therapy”[Mesh] OR “Recovery of Function”[Mesh] OR “Occupational Therapy”[Mesh] OR “Allied Health Occupations”[Mesh] OR “Therapeutics”[Mesh] OR “Physical Therapist Assistants”[Mesh] OR “Allied Health Personnel”[Mesh] OR “Physical Therapy Modalities”[Mesh] OR “Physical Therapy Specialty”[Mesh]))

###### Free terms

Physiotherapy OR “physical rehabilitation” OR “physical therapy” OR “physical therapy modalities” OR “physiotherapy techniques” OR “physiotherapy rehabilitation” OR “physical therapist” OR physiotherapist OR “occupational therapy” OR “allied health worker” OR “allied health professional” OR “occupational therapist” OR nurse OR “rehabilitation centre” OR rehabilitation OR rehabilitative OR neurorehabilitation OR “neurological physiotherapy” OR “neurological rehabilitation” OR “Neurologic Rehabilitation” OR “stroke rehabilitation” OR mobilization OR “activities of daily living” OR “functional activities” OR “functional training” OR “exercise therapy” OR mobility OR “functional assessment” OR “Disability Evaluation” OR “virtual rehabilitation” OR telerehabilitation OR “geriatric rehabilitation” OR “recreational therapy” OR “vocational rehabilitation”

###### Mesh terms + Free terms

((“Rehabilitation”[Mesh] OR “Physical and Rehabilitation Medicine”[Mesh] OR “Stroke Rehabilitation”[Mesh] OR “Neurological Rehabilitation”[Mesh] OR “Orthopedic Procedures”[Mesh] OR “Telerehabilitation”[Mesh] OR “Exercise Therapy”[Mesh] OR “Treatment Outcome”[Mesh] OR “Rehabilitation Centers”[Mesh] OR “rehabilitation” [Subheading] OR “Activities of Daily Living”[Mesh] OR “Recreation Therapy”[Mesh] OR “Recovery of Function”[Mesh] OR “Occupational Therapy”[Mesh] OR “Allied Health Occupations”[Mesh] OR “Therapeutics”[Mesh] OR “Physical Therapist Assistants”[Mesh] OR “Allied Health Personnel”[Mesh] OR “Physical Therapy Modalities”[Mesh] OR “Physical Therapy Specialty”[Mesh])) OR (Physiotherapy OR “physical rehabilitation” OR “physical therapy” OR “physical therapy modalities” OR “physiotherapy techniques” OR “physiotherapy rehabilitation” OR “physical therapist” OR physiotherapist OR “occupational therapy” OR “allied health worker” OR “allied health professional” OR “occupational therapist” OR nurse OR “rehabilitation centre” OR rehabilitation OR rehabilitative OR neurorehabilitation OR “neurological physiotherapy” OR “neurological rehabilitation” OR “Neurologic Rehabilitation” OR “stroke rehabilitation” OR mobilization OR “activities of daily living” OR “functional activities” OR “functional training” OR “exercise therapy” OR mobility OR “functional assessment” OR “Disability Evaluation” OR “virtual rehabilitation” OR telerehabilitation OR “geriatric rehabilitation” OR “recreational therapy” OR “vocational rehabilitation”)

##### Descriptor 3: Technology

12,167,954 results

###### Mesh terms

(“Virtual Reality”[Mesh] OR “Virtual Reality Exposure Therapy”[Mesh] OR “Robotics”[Mesh] OR “Telerehabilitation”[Mesh] OR “Video Games”[Mesh] OR “Technology”[Mesh] OR “Remote Sensing Technology”[Mesh] OR “Wireless Technology”[Mesh] OR “Biomedical Technology”[Mesh] OR “Electronics”[Mesh] OR “Electronic Supplementary Materials” [Publication Type] OR “Mobile Applications”[Mesh] OR “Videodisc Recording”[Mesh] OR “Electrical Equipment and Supplies”[Mesh] OR “Telemedicine”[Mesh] OR “Remote Consultation”[Mesh])

###### Free terms

((Telemedicine OR telemetry OR videoconferencing OR telecommunications OR “computer communication networks” OR “remote consultation” OR “remote sensing technology” OR “virtual Rehabilitation” OR “Virtual Rehabilitations” OR telerehabilitation OR tele-rehabilitation OR telerehab OR telehealth OR tele-health OR telehomecare OR tele-homecare OR telecoaching OR tele-coaching OR telecommunication* OR “Remote Rehabilitation” OR videoconference* OR video-conferenc* OR videoconsultation OR video-consultation OR telestroke OR teleconference* OR tele-conference* OR teleconsultation OR tele-consultation OR telecare OR ehealth OR e-health OR telepractice OR teletherap* OR telehealth OR mhealth OR m-health OR “m health” OR “mobile health” OR remote* OR distance* OR distant OR “occupational therap*”) OR Computer OR microcomputer OR minicomputer OR “cell phone” OR “mobile application” OR telephone* OR phone* OR cell* OR smart* OR mobile OR android OR video* OR internet* OR computer* OR sensor* OR modem OR webcam OR website* OR email OR web OR comput* OR device OR app OR smartphone OR text-messag* OR tablet OR technolog* OR telephone OR “electronic mail” OR internet OR “virtual reality” OR “virtual environment*” OR “Educational Virtual Realit*”) OR robotics OR automation OR orthotic devices OR Robot-assisted OR “equipment and supplies” OR Telerobotics OR “self-help devices” OR “physical therapy modalities” OR “occupational therapy” OR “therapy, computer-assisted” OR “man-machine systems” OR robot* OR orthos* OR orthotic OR automat* OR “computer aided” OR “computer assisted” OR electromechanical OR electro-mechanical OR mechanical OR mechanised OR mechanized

###### Mesh terms + Free terms

((“Virtual Reality”[Mesh] OR “Virtual Reality Exposure Therapy”[Mesh] OR “Robotics”[Mesh] OR “Telerehabilitation”[Mesh] OR “Video Games”[Mesh] OR “Technology”[Mesh] OR “Remote Sensing Technology”[Mesh] OR “Wireless Technology”[Mesh] OR “Biomedical Technology”[Mesh] OR “Electronics”[Mesh] OR “Electronic Supplementary Materials”

[Publication Type] OR “Mobile Applications”[Mesh] OR “Videodisc Recording”[Mesh] OR “Electrical Equipment and Supplies”[Mesh] OR “Telemedicine”[Mesh] OR “Remote Consultation”[Mesh])) OR (((Telemedicine OR telemetry OR videoconferencing OR telecommunications OR “computer communication networks” OR “remote consultation” OR “remote sensing technology” OR “virtual Rehabilitation” OR “Virtual Rehabilitations” OR telerehabilitation OR tele-rehabilitation OR telerehab OR telehealth OR tele-health OR telehomecare OR tele-homecare OR telecoaching OR tele-coaching OR telecommunication* OR “Remote Rehabilitation” OR videoconference* OR video-conferenc* OR videoconsultation OR video-consultation OR telestroke OR teleconference* OR tele-conference* OR teleconsultation OR tele-consultation OR telecare OR ehealth OR e-health OR telepractice OR teletherap* OR telehealth OR mhealth OR m-health OR “m health” OR “mobile health” OR remote* OR distance* OR distant OR “occupational therap*”) OR Computer OR microcomputer OR minicomputer OR “cell phone” OR “mobile application” OR telephone* OR phone* OR cell* OR smart* OR mobile OR android OR video* OR internet* OR computer* OR sensor* OR modem OR webcam OR website* OR email OR web OR comput* OR device OR app OR smartphone OR text-messag* OR tablet OR technolog* OR telephone OR “electronic mail” OR internet OR “virtual reality” OR “virtual environment*” OR “Educational Virtual Realit*”) OR robotics OR automation OR orthotic devices OR Robot-assisted OR “equipment and supplies” OR Telerobotics OR “self-help devices” OR “physical therapy modalities” OR “occupational therapy” OR “therapy, computer-assisted” OR “man-machine systems” OR robot* OR orthos* OR orthotic OR automat* OR “computer aided” OR “computer assisted” OR electromechanical OR electro-mechanical OR mechanical OR mechanised OR mechanized)

##### Descriptor 4: Low- and-middle income countries

Low and middle income countries according to the World Bank, accessed at the following link: https://datahelpdesk.worldbank.org/knowledgebase/articles/906519-4,829,387 results

###### Mesh terms

(((((((((((((((((((((((((((((((((((((((((((((((((((((((((((((((((((((((((((((((((((((((((((((((((((((((((((((((((((((((((((((((((((((“Developing Countries”[Mesh]) OR “Afghanistan”[Mesh]) OR “Benin”[Mesh]) OR “Burkina Faso”[Mesh])) OR “Burundi”[Mesh]) OR “Central African Republic”[Mesh]) OR “Chad”[Mesh]) OR “Democratic Republic of the Congo”[Mesh]) OR “Ethiopia”[Mesh]) OR “Gambia”[Mesh]) OR “Guinea”[Mesh]) OR “Haiti”[Mesh]) OR “Democratic People’s Republic of Korea”[Mesh]) OR “Liberia”[Mesh]) OR “Madagascar”[Mesh]) OR “Malawi”[Mesh]) OR “Nepal”[Mesh]) OR “Niger”[Mesh]) OR “Rwanda”[Mesh]) OR “Sierra Leone”[Mesh]) OR “Somalia”[Mesh]) OR “South Sudan”[Mesh]) OR “Syria”[Mesh]) OR “Tajikistan”[Mesh]) OR “Tanzania”[Mesh]) OR “Togo”[Mesh]) OR “Uganda”[Mesh]) OR “Yemen”[Mesh]) OR “Mali”[Mesh]) OR “Angola”[Mesh]) OR “Bangladesh”[Mesh]) OR “Bhutan”[Mesh]) OR “Bolivia”[Mesh]) OR “Cabo Verde”[Mesh]) OR “Cambodia”[Mesh]) OR “Cameroon”[Mesh]) OR “Comoros”[Mesh]) OR “Cote d’Ivoire”[Mesh]) OR “Djibouti”[Mesh]) OR “Egypt”[Mesh]) OR “El Salvador”[Mesh]) OR “Ghana”[Mesh]) OR “Honduras”[Mesh]) OR “India”[Mesh]) OR “Papua New Guinea”[Mesh]) OR “Indonesia”[Mesh]) OR “Kenya”[Mesh]) OR “Micronesia”[Mesh]) OR “Kyrgyzstan”[Mesh]) OR “Laos”[Mesh]) OR “Lesotho”[Mesh]) OR “Mauritania”[Mesh]) OR “Moldova”[Mesh]) OR “Mongolia”[Mesh]) OR “Morocco”[Mesh]) OR “Myanmar”[Mesh]) OR “Nicaragua”[Mesh]) OR “Nigeria”[Mesh]) OR “Pakistan”[Mesh]) OR “Philippines”[Mesh]) OR “Sao Tome and Principe”[Mesh]) OR “Senegal”[Mesh]) OR “Melanesia”[Mesh]) OR “Sudan”[Mesh]) OR “Eswatini”[Mesh]) OR “Timor-Leste”[Mesh]) OR “Tunisia”[Mesh]) OR “Ukraine”[Mesh]) OR “Uzbekistan”[Mesh]) OR “Vanuatu”[Mesh]) OR “Vietnam”[Mesh]) OR “Zambia”[Mesh]) OR “Zimbabwe”[Mesh]) OR “Albania”[Mesh]) OR “Algeria”[Mesh]) OR “American Samoa”[Mesh]) OR “Argentina”[Mesh]) OR “Armenia”[Mesh]) OR “Azerbaijan”[Mesh]) OR “Republic of Belarus”[Mesh]) OR “Belize”[Mesh]) OR “Bosnia and Herzegovina”[Mesh]) OR “Botswana”[Mesh]) OR “Brazil”[Mesh]) OR “Bulgaria”[Mesh]) OR “China”[Mesh]) OR “Colombia”[Mesh]) OR “Costa Rica”[Mesh]) OR “Cuba”[Mesh]) OR “Dominica”[Mesh]) OR “Dominican Republic”[Mesh]) OR “Equatorial Guinea”[Mesh]) OR “Ecuador”[Mesh]) OR “Fiji”[Mesh]) OR “Gabon”[Mesh]) OR “Georgia (Republic)”[Mesh]) OR “Grenada”[Mesh]) OR “Guatemala”[Mesh]) OR “Guyana”[Mesh]) OR “Iran”[Mesh]) OR “Iraq”[Mesh]) OR “Jamaica”[Mesh]) OR “Jordan”[Mesh]) OR “Kazakhstan”[Mesh]) OR “Kosovo”[Mesh]) OR “Lebanon”[Mesh]) OR “Libya”[Mesh]) OR “Republic of North Macedonia”[Mesh]) OR “Malaysia”[Mesh]) OR “Indian Ocean Islands”[Mesh]) OR “Mexico”[Mesh]) OR “Montenegro”[Mesh]) OR “Namibia”[Mesh]) OR “Paraguay”[Mesh]) OR “Peru”[Mesh]) OR “Russia”[Mesh]) OR “Samoa”[Mesh]) OR “Serbia”[Mesh]) OR “Sri Lanka”[Mesh]) OR “South Africa”[Mesh]) OR “Saint Lucia”[Mesh]) OR “Saint Vincent and the Grenadines”[Mesh]) OR “Suriname”[Mesh]) OR “Thailand”[Mesh]) OR “Tonga”[Mesh]) OR “Turkey”[Mesh]) OR “Turkmenistan”[Mesh]) OR “Venezuela”[Mesh]) OR “Mozambique”[Mesh]) OR “Eritrea”[Mesh]) OR “Guinea-Bissau”[Mesh]) OR “Congo”[Mesh])

###### Free terms

(“lower-middle income country” OR “lower-middle income countries” OR “lower-middle income nations” OR “lower-middle income nation” OR “low income country” OR “low-income countries” OR “low-income nation” OR “low-income nations” OR “middle income countries” OR “lower income countries” OR “middle income country” OR “middle-income countries” OR “middle income nation” OR “middle-income nation” OR “upper-middle income country” OR “upper-middle income countries” OR “upper-middle income nations” OR “upper-middle income nation” OR “developing nation” OR “developing nations” OR “underdeveloped nation” OR “underdeveloped nations” OR “developing countries” OR “developing country” OR “third world” OR “resource constrained” OR “low resource setting” OR “low resource” OR “limited resource” OR “less resourced” OR “resource poor” OR “resource limited”) OR “Afghanistan” OR “Benin” OR “Burkina Faso” OR “Burundi” OR “Central African Republic” OR “Chad” OR “Democratic Republic of the Congo” OR “Ethiopia” OR “Gambia” OR “Guinea” OR “Haiti” OR “Democratic People’s Republic of Korea” OR “Liberia” OR “Madagascar” OR “Malawi” OR “Nepal” OR “Niger” OR “Rwanda” OR “Sierra Leone” OR “Somalia” OR “South Sudan” OR “Syria” OR “Tajikistan” OR “Tanzania” OR “Togo” OR “Uganda” OR “Yemen” OR “Mali” OR “Angola” OR “Bangladesh” OR “Bhutan” OR “Bolivia” OR “Cabo Verde” OR “Cambodia” OR “Cameroon” OR “Comoros” OR “Cote d’Ivoire” OR “Djibouti” OR “Egypt” OR “El Salvador” OR “Ghana” OR “Honduras” OR “India” OR “Papua New Guinea” OR “Indonesia” OR “Kenya” OR “Micronesia” OR “Kyrgyzstan” OR “Laos” OR “Lesotho” OR “Mauritania” OR “Moldova” OR “Mongolia” OR “Morocco” OR “Myanmar” OR “Nicaragua” OR “Nigeria” OR “Pakistan” OR “Philippines” OR “Sao Tome and Principe” OR “Senegal” OR “Melanesia” OR “Sudan” OR “Eswatini” OR “Timor-Leste” OR “Tunisia” OR “Ukraine” OR “Uzbekistan” OR “Vanuatu” OR “Vietnam” OR “Zambia” OR “Zimbabwe” OR “Albania” OR “Algeria” OR “American Samoa” OR “Argentina” OR “Armenia” OR “Azerbaijan” OR “Republic of Belarus” OR “Belize” OR “Bosnia and Herzegovina” OR “Botswana” OR “Brazil” OR “Bulgaria” OR “China” OR “Colombia” OR “Costa Rica” OR “Cuba” OR “Dominica” OR “Dominican Republic” OR “Equatorial Guinea” OR “Ecuador” OR “Fiji” OR “Gabon” OR “Georgia (Republic)” OR “Grenada” OR “Guatemala” OR “Guyana” OR “Iran” OR “Iraq” OR “Jamaica” OR “Jordan” OR “Kazakhstan” OR “Kosovo” OR “Lebanon” OR “Libya” OR “Republic of North Macedonia” OR “Malaysia” OR “Indian Ocean Islands” OR “Mexico” OR “Montenegro” OR “Namibia” OR “Paraguay” OR “Peru” OR “Russia” OR “Samoa” OR “Serbia” OR “Sri Lanka” OR “South Africa” OR “Saint Lucia” OR “Saint Vincent and the Grenadines” OR “Suriname” OR “Thailand” OR “Tonga” OR “Turkey” OR “Turkmenistan” OR “Venezuela” OR “Mozambique” OR “Eritrea” OR “Guinea-Bissau” OR “Congo”

###### Mesh terms + Free terms

(((((((((((((((((((((((((((((((((((((((((((((((((((((((((((((((((((((((((((((((((((((((((((((((((((((((((((((((((((((((((((((((((((((((“Developing Countries”[Mesh]) OR “Afghanistan”[Mesh]) OR “Benin”[Mesh]) OR “Burkina Faso”[Mesh])) OR “Burundi”[Mesh]) OR “Central African Republic”[Mesh]) OR “Chad”[Mesh]) OR “Democratic Republic of the Congo”[Mesh]) OR “Ethiopia”[Mesh]) OR “Gambia”[Mesh]) OR “Guinea”[Mesh]) OR “Haiti”[Mesh]) OR “Democratic People’s Republic of Korea”[Mesh]) OR “Liberia”[Mesh]) OR “Madagascar”[Mesh]) OR “Malawi”[Mesh]) OR “Nepal”[Mesh]) OR “Niger”[Mesh]) OR “Rwanda”[Mesh]) OR “Sierra Leone”[Mesh]) OR “Somalia”[Mesh]) OR “South Sudan”[Mesh]) OR “Syria”[Mesh]) OR “Tajikistan”[Mesh]) OR “Tanzania”[Mesh]) OR “Togo”[Mesh]) OR “Uganda”[Mesh]) OR “Yemen”[Mesh]) OR “Mali”[Mesh]) OR “Angola”[Mesh]) OR “Bangladesh”[Mesh]) OR “Bhutan”[Mesh]) OR “Bolivia”[Mesh]) OR “Cabo Verde”[Mesh]) OR “Cambodia”[Mesh]) OR “Cameroon”[Mesh]) OR “Comoros”[Mesh]) OR “Cote d’Ivoire”[Mesh]) OR “Djibouti”[Mesh]) OR “Egypt”[Mesh]) OR “El Salvador”[Mesh]) OR “Ghana”[Mesh]) OR “Honduras”[Mesh]) OR “India”[Mesh]) OR “Papua New Guinea”[Mesh]) OR “Indonesia”[Mesh]) OR “Kenya”[Mesh]) OR “Micronesia”[Mesh]) OR “Kyrgyzstan”[Mesh]) OR “Laos”[Mesh]) OR “Lesotho”[Mesh]) OR “Mauritania”[Mesh]) OR “Moldova”[Mesh]) OR “Mongolia”[Mesh]) OR “Morocco”[Mesh]) OR “Myanmar”[Mesh]) OR “Nicaragua”[Mesh]) OR “Nigeria”[Mesh]) OR “Pakistan”[Mesh]) OR “Philippines”[Mesh]) OR “Sao Tome and Principe”[Mesh]) OR “Senegal”[Mesh]) OR “Melanesia”[Mesh]) OR “Sudan”[Mesh]) OR “Eswatini”[Mesh]) OR “Timor-Leste”[Mesh]) OR “Tunisia”[Mesh]) OR “Ukraine”[Mesh]) OR “Uzbekistan”[Mesh]) OR “Vanuatu”[Mesh]) OR “Vietnam”[Mesh]) OR “Zambia”[Mesh]) OR “Zimbabwe”[Mesh]) OR “Albania”[Mesh]) OR “Algeria”[Mesh]) OR “American Samoa”[Mesh]) OR “Argentina”[Mesh]) OR “Armenia”[Mesh]) OR “Azerbaijan”[Mesh]) OR “Republic of Belarus”[Mesh]) OR “Belize”[Mesh]) OR “Bosnia and Herzegovina”[Mesh]) OR “Botswana”[Mesh]) OR “Brazil”[Mesh]) OR “Bulgaria”[Mesh]) OR “China”[Mesh]) OR “Colombia”[Mesh]) OR “Costa Rica”[Mesh]) OR “Cuba”[Mesh]) OR “Dominica”[Mesh]) OR “Dominican Republic”[Mesh]) OR “Equatorial Guinea”[Mesh]) OR “Ecuador”[Mesh]) OR “Fiji”[Mesh]) OR “Gabon”[Mesh]) OR “Georgia (Republic)”[Mesh]) OR “Grenada”[Mesh]) OR “Guatemala”[Mesh]) OR “Guyana”[Mesh]) OR “Iran”[Mesh]) OR “Iraq”[Mesh]) OR “Jamaica”[Mesh]) OR “Jordan”[Mesh]) OR “Kazakhstan”[Mesh]) OR “Kosovo”[Mesh]) OR “Lebanon”[Mesh]) OR “Libya”[Mesh]) OR “Republic of North Macedonia”[Mesh]) OR “Malaysia”[Mesh]) OR “Indian Ocean Islands”[Mesh]) OR “Mexico”[Mesh]) OR “Montenegro”[Mesh]) OR “Namibia”[Mesh]) OR “Paraguay”[Mesh]) OR “Peru”[Mesh]) OR “Russia”[Mesh]) OR “Samoa”[Mesh]) OR “Serbia”[Mesh]) OR “Sri Lanka”[Mesh]) OR “South Africa”[Mesh]) OR “Saint Lucia”[Mesh]) OR “Saint Vincent and the Grenadines”[Mesh]) OR “Suriname”[Mesh]) OR “Thailand”[Mesh]) OR “Tonga”[Mesh]) OR “Turkey”[Mesh]) OR “Turkmenistan”[Mesh]) OR “Venezuela”[Mesh]) OR “Mozambique”[Mesh]) OR “Eritrea”[Mesh]) OR “Guinea-Bissau”[Mesh]) OR “Congo”[Mesh])) OR (“lower-middle income country” OR “lower-middle income countries” OR “lower-middle income nations” OR “lower-middle income nation” OR “low income country” OR “low-income countries” OR “low-income nation” OR “low-income nations” OR “middle income countries” OR “lower income countries” OR “middle income country” OR “middle-income countries” OR “middle income nation” OR “middle-income nation” OR “upper-middle income country” OR “upper-middle income countries” OR “upper-middle income nations” OR “upper-middle income nation” OR “developing nation” OR “developing nations” OR “underdeveloped nation” OR “underdeveloped nations” OR “developing countries” OR “developing country” OR “third world” OR “resource constrained” OR “low resource setting” OR “low resource” OR “limited resource” OR “less resourced” OR “resource poor” OR “resource limited”)) OR (“Afghanistan” OR “Benin” OR “Burkina Faso” OR “Burundi” OR “Central African Republic” OR “Chad” OR “Democratic Republic of the Congo” OR “Ethiopia” OR “Gambia” OR “Guinea” OR “Haiti” OR “Democratic People’s Republic of Korea” OR “Liberia” OR “Madagascar” OR “Malawi” OR “Nepal” OR “Niger” OR “Rwanda” OR “Sierra Leone” OR “Somalia” OR “South Sudan” OR “Syria” OR “Tajikistan” OR “Tanzania” OR “Togo” OR “Uganda” OR “Yemen” OR “Mali” OR “Angola” OR “Bangladesh” OR “Bhutan” OR “Bolivia” OR “Cabo Verde” OR “Cambodia” OR “Cameroon” OR “Comoros” OR “Cote d’Ivoire” OR “Djibouti” OR “Egypt” OR “El Salvador” OR “Ghana” OR “Honduras” OR “India” OR “Papua New Guinea” OR “Indonesia” OR “Kenya” OR “Micronesia” OR “Kyrgyzstan” OR “Laos” OR “Lesotho” OR “Mauritania” OR “Moldova” OR “Mongolia” OR “Morocco” OR “Myanmar” OR “Nicaragua” OR “Nigeria” OR “Pakistan” OR “Philippines” OR “Sao Tome and Principe” OR “Senegal” OR “Melanesia” OR “Sudan” OR “Eswatini” OR “Timor-Leste” OR “Tunisia” OR “Ukraine” OR “Uzbekistan” OR “Vanuatu” OR “Vietnam” OR “Zambia” OR “Zimbabwe” OR “Albania” OR “Algeria” OR “American Samoa” OR “Argentina” OR “Armenia” OR “Azerbaijan” OR “Republic of Belarus” OR “Belize” OR “Bosnia and Herzegovina” OR “Botswana” OR “Brazil” OR “Bulgaria” OR “China” OR “Colombia” OR “Costa Rica” OR “Cuba” OR “Dominica” OR “Dominican Republic” OR “Equatorial Guinea” OR “Ecuador” OR “Fiji” OR “Gabon” OR “Georgia (Republic)” OR “Grenada” OR “Guatemala” OR “Guyana” OR “Iran” OR “Iraq” OR “Jamaica” OR “Jordan” OR “Kazakhstan” OR “Kosovo” OR “Lebanon” OR “Libya” OR “Republic of North Macedonia” OR “Malaysia” OR “Indian Ocean Islands” OR “Mexico” OR “Montenegro” OR “Namibia” OR “Paraguay” OR “Peru” OR “Russia” OR “Samoa” OR “Serbia” OR “Sri Lanka” OR “South Africa” OR “Saint Lucia” OR “Saint Vincent and the Grenadines” OR “Suriname” OR “Thailand” OR “Tonga” OR “Turkey” OR “Turkmenistan” OR “Venezuela” OR “Mozambique” OR “Eritrea” OR “Guinea-Bissau” OR “Congo”)

##### Descriptor 5: Randomized controlled trials (RCTs)

930,517 results

###### Mesh terms

(((“Randomized Controlled Trial” [Publication Type] OR “Clinical Trial” [Publication Type] OR “Controlled Clinical Trial” [Publication Type]

###### Free terms

“randomized controlled trial” OR rct OR “controlled trial”

###### Mesh terms + Free terms

“Randomized Controlled Trial” [Publication Type] OR “Clinical Trial” [Publication Type] OR “Controlled Clinical Trial” [Publication Type] OR “randomized controlled trial” OR rct OR “controlled trial”

###### All (Mesh terms + Free terms)

Filters applied: English, French, Portuguese, Spanish. 1476 results ((((((((“Stroke”[Mesh] OR “Stroke, Lacunar”[Mesh] OR “Infarction, Posterior Cerebral Artery”[Mesh] OR “Brain Stem Infarctions”[Mesh] OR “Infarction, Middle Cerebral Artery”[Mesh] OR “Infarction, Anterior Cerebral Artery”[Mesh] OR “Cerebral Hemorrhage”[Mesh] OR “Brain Ischemia”[Mesh]) OR “Paresis”[Mesh]) OR “Hemiplegia” [Mesh])) OR (“Stroke” [Majr] OR “stroke*” OR “strokes” OR “Brain Infarction” OR “brain infarctions” OR “cerebrovascular disorder” OR “cerebrovascular disorders” OR “cerebrovascular accident” OR “cerebrovascular accidents” OR “cva” OR “cvas” OR “cerebral infarction” OR “cerebral infarctions” OR “Intracranial Embolism and Thrombosis” OR “cerebrovascular disease” OR “cerebrovascular diseases” OR “cerebral haemorrhage” OR “cerebral hemorrhage” OR “brain ischemia” OR “brain ischaemia” OR “cerebral ischemia” OR “cerebral ischaemia” OR “brain vascular accident” OR “cerebrovascular Stroke” OR “Cerebral Stroke” OR “Acute Stroke” OR “Acute Strokes” OR “basal ganglia cerebrovascular disease” OR “intracranial arterial diseases” OR “intracranial embolism and thrombosis” OR “intracranial hemorrhages” OR “poststroke” OR “apoplex*” OR “cerebral vasc*” OR “brain vasc*” OR “Paresis” OR “Pareses” OR “Muscular Paresis” OR “Muscular Pareses” OR “Muscle Paresis” OR “Muscle Pareses” OR “Hemiparesis” OR “Hemipareses” OR “hemiplegia” OR “hemipleg*”OR “hemipar*” OR “paretic*”)) **AND** (((“Rehabilitation”[Mesh] OR “Physical and Rehabilitation Medicine”[Mesh] OR “Stroke Rehabilitation”[Mesh] OR “Neurological Rehabilitation”[Mesh] OR “Orthopedic Procedures”[Mesh] OR “Telerehabilitation”[Mesh] OR “Exercise Therapy”[Mesh] OR “Treatment Outcome”[Mesh] OR “Rehabilitation Centers”[Mesh] OR “rehabilitation” [Subheading] OR “Activities of Daily Living”[Mesh] OR “Recreation Therapy”[Mesh] OR “Recovery of Function”[Mesh] OR “Occupational Therapy”[Mesh] OR “Allied Health Occupations”[Mesh] OR “Therapeutics”[Mesh] OR “Physical Therapist Assistants”[Mesh] OR “Allied Health Personnel”[Mesh] OR “Physical Therapy Modalities”[Mesh] OR “Physical Therapy Specialty”[Mesh])) OR (Physiotherapy OR “physical rehabilitation” OR “physical therapy” OR “physical therapy modalities” OR “physiotherapy techniques” OR “physiotherapy rehabilitation” OR “physical therapist” OR physiotherapist OR “occupational therapy” OR “allied health worker” OR “allied health professional” OR “occupational therapist” OR nurse OR “rehabilitation centre” OR rehabilitation OR rehabilitative OR neurorehabilitation OR “neurological physiotherapy” OR “neurological rehabilitation” OR “Neurologic Rehabilitation” OR “stroke rehabilitation” OR mobilization OR “activities of daily living” OR “functional activities” OR “functional training” OR “exercise therapy” OR mobility OR “functional assessment” OR “Disability Evaluation” OR “virtual rehabilitation” OR telerehabilitation OR “geriatric rehabilitation” OR “recreational therapy” OR “vocational rehabilitation”))) **AND** (((“Virtual Reality”[Mesh] OR “Virtual Reality Exposure Therapy”[Mesh] OR “Robotics”[Mesh] OR “Telerehabilitation”[Mesh] OR “Video Games”[Mesh] OR “Technology”[Mesh] OR “Remote Sensing Technology”[Mesh] OR “Wireless Technology”[Mesh] OR “Biomedical Technology”[Mesh] OR “Electronics”[Mesh] OR “Electronic Supplementary Materials” [Publication Type] OR “Mobile Applications”[Mesh] OR “Videodisc Recording”[Mesh] OR “Electrical Equipment and Supplies”[Mesh] OR “Telemedicine”[Mesh] OR “Remote Consultation”[Mesh])) OR (((Telemedicine OR telemetry OR videoconferencing OR telecommunications OR “computer communication networks” OR “remote consultation” OR “remote sensing technology” OR “virtual Rehabilitation” OR “Virtual Rehabilitations” OR telerehabilitation OR tele-rehabilitation OR telerehab OR telehealth OR tele-health OR telehomecare OR tele-homecare OR telecoaching OR tele-coaching OR telecommunication* OR “Remote Rehabilitation” OR videoconference* OR video-conferenc* OR videoconsultation OR video-consultation OR telestroke OR teleconference* OR tele-conference* OR teleconsultation OR tele-consultation OR telecare OR ehealth OR e-health OR telepractice OR teletherap* OR telehealth OR mhealth OR m-health OR “m health” OR “mobile health” OR remote* OR distance* OR distant OR “occupational therap*”) OR Computer OR microcomputer OR minicomputer OR “cell phone” OR “mobile application” OR telephone* OR phone* OR cell* OR smart* OR mobile OR android OR video* OR internet* OR computer* OR sensor* OR modem OR webcam OR website* OR email OR web OR comput* OR device OR app OR smartphone OR text-messag* OR tablet OR technolog* OR telephone OR “electronic mail” OR internet OR “virtual reality” OR “virtual environment*” OR “Educational Virtual Realit*”) OR robotics OR automation OR orthotic devices OR Robot-assisted OR “equipment and supplies” OR Telerobotics OR “self-help devices” OR “physical therapy modalities” OR “occupational therapy” OR “therapy, computer-assisted” OR “man-machine systems” OR robot* OR orthos* OR orthotic OR automat* OR “computer aided” OR “computer assisted” OR electromechanical OR electro-mechanical OR mechanical OR mechanised OR mechanized))) **AND** ((((((((((((((((((((((((((((((((((((((((((((((((((((((((((((((((((((((((((((((((((((((((((((((((((((((((((((((((((((((((((((((((((((((((“Developing Countries”[Mesh]) OR “Afghanistan”[Mesh]) OR “Benin”[Mesh]) OR “Burkina Faso”[Mesh])) OR “Burundi”[Mesh]) OR “Central African Republic”[Mesh]) OR “Chad”[Mesh]) OR “Democratic Republic of the Congo”[Mesh]) OR “Ethiopia”[Mesh]) OR “Gambia”[Mesh]) OR “Guinea”[Mesh]) OR “Haiti”[Mesh]) OR “Democratic People’s Republic of Korea”[Mesh]) OR “Liberia”[Mesh]) OR “Madagascar”[Mesh]) OR “Malawi”[Mesh]) OR “Nepal”[Mesh]) OR “Niger”[Mesh]) OR “Rwanda”[Mesh]) OR “Sierra Leone”[Mesh]) OR “Somalia”[Mesh]) OR “South Sudan”[Mesh]) OR “Syria”[Mesh]) OR “Tajikistan”[Mesh]) OR “Tanzania”[Mesh]) OR “Togo”[Mesh]) OR “Uganda”[Mesh]) OR “Yemen”[Mesh]) OR “Mali”[Mesh]) OR “Angola”[Mesh]) OR “Bangladesh”[Mesh]) OR “Bhutan”[Mesh]) OR “Bolivia”[Mesh]) OR “Cabo Verde”[Mesh]) OR “Cambodia”[Mesh]) OR “Cameroon”[Mesh]) OR “Comoros”[Mesh]) OR “Cote d’Ivoire”[Mesh]) OR “Djibouti”[Mesh]) OR “Egypt”[Mesh]) OR “El Salvador”[Mesh]) OR “Ghana”[Mesh]) OR “Honduras”[Mesh]) OR “India”[Mesh]) OR “Papua New Guinea”[Mesh]) OR “Indonesia”[Mesh]) OR “Kenya”[Mesh]) OR “Micronesia”[Mesh]) OR “Kyrgyzstan”[Mesh]) OR “Laos”[Mesh]) OR “Lesotho”[Mesh]) OR “Mauritania”[Mesh]) OR “Moldova”[Mesh]) OR “Mongolia”[Mesh]) OR “Morocco”[Mesh]) OR “Myanmar”[Mesh]) OR “Nicaragua”[Mesh]) OR “Nigeria”[Mesh]) OR “Pakistan”[Mesh]) OR “Philippines”[Mesh]) OR “Sao Tome and Principe”[Mesh]) OR “Senegal”[Mesh]) OR “Melanesia”[Mesh]) OR “Sudan”[Mesh]) OR “Eswatini”[Mesh]) OR “Timor-Leste”[Mesh]) OR “Tunisia”[Mesh]) OR “Ukraine”[Mesh]) OR “Uzbekistan”[Mesh]) OR “Vanuatu”[Mesh]) OR “Vietnam”[Mesh]) OR “Zambia”[Mesh]) OR “Zimbabwe”[Mesh]) OR “Albania”[Mesh]) OR “Algeria”[Mesh]) OR “American Samoa”[Mesh]) OR “Argentina”[Mesh]) OR “Armenia”[Mesh])

OR “Azerbaijan”[Mesh]) OR “Republic of Belarus”[Mesh]) OR “Belize”[Mesh]) OR “Bosnia and Herzegovina”[Mesh]) OR “Botswana”[Mesh]) OR “Brazil”[Mesh]) OR “Bulgaria”[Mesh]) OR “China”[Mesh]) OR “Colombia”[Mesh]) OR “Costa Rica”[Mesh]) OR “Cuba”[Mesh]) OR “Dominica”[Mesh]) OR “Dominican Republic”[Mesh]) OR “Equatorial Guinea”[Mesh]) OR “Ecuador”[Mesh]) OR “Fiji”[Mesh]) OR “Gabon”[Mesh]) OR “Georgia (Republic)”[Mesh]) OR “Grenada”[Mesh]) OR “Guatemala”[Mesh]) OR “Guyana”[Mesh]) OR “Iran”[Mesh]) OR “Iraq”[Mesh]) OR “Jamaica”[Mesh]) OR “Jordan”[Mesh]) OR “Kazakhstan”[Mesh]) OR “Kosovo”[Mesh]) OR “Lebanon”[Mesh]) OR “Libya”[Mesh]) OR “Republic of North Macedonia”[Mesh]) OR “Malaysia”[Mesh]) OR “Indian Ocean Islands”[Mesh]) OR “Mexico”[Mesh]) OR “Montenegro”[Mesh]) OR “Namibia”[Mesh]) OR “Paraguay”[Mesh]) OR “Peru”[Mesh]) OR “Russia”[Mesh]) OR “Samoa”[Mesh]) OR “Serbia”[Mesh]) OR “Sri Lanka”[Mesh]) OR “South Africa”[Mesh]) OR “Saint Lucia”[Mesh]) OR “Saint Vincent and the Grenadines”[Mesh]) OR “Suriname”[Mesh]) OR “Thailand”[Mesh]) OR “Tonga”[Mesh]) OR “Turkey”[Mesh]) OR “Turkmenistan”[Mesh]) OR “Venezuela”[Mesh]) OR “Mozambique”[Mesh]) OR “Eritrea”[Mesh]) OR “Guinea-Bissau”[Mesh]) OR “Congo”[Mesh])) OR (“lower-middle income country” OR “lower-middle income countries” OR “lower-middle income nations” OR “lower-middle income nation” OR “low income country” OR “low-income countries” OR “low-income nation” OR “low-income nations” OR “middle income countries” OR “lower income countries” OR “middle income country” OR “middle-income countries” OR “middle income nation” OR “middle-income nation” OR “upper-middle income country” OR “upper-middle income countries” OR “upper-middle income nations” OR “upper-middle income nation” OR “developing nation” OR “developing nations” OR “underdeveloped nation” OR “underdeveloped nations” OR “developing countries” OR “developing country” OR “third world” OR “resource constrained” OR “low resource setting” OR “low resource” OR “limited resource” OR “less resourced” OR “resource poor” OR “resource limited”)) OR (“Afghanistan” OR “Benin” OR “Burkina Faso” OR “Burundi” OR “Central African Republic” OR “Chad” OR “Democratic Republic of the Congo” OR “Ethiopia” OR “Gambia” OR “Guinea” OR “Haiti” OR “Democratic People’s Republic of Korea” OR “Liberia” OR “Madagascar” OR “Malawi” OR “Nepal” OR “Niger” OR “Rwanda” OR “Sierra Leone” OR “Somalia” OR “South Sudan” OR “Syria” OR “Tajikistan” OR “Tanzania” OR “Togo” OR “Uganda” OR “Yemen” OR “Mali” OR “Angola” OR “Bangladesh” OR “Bhutan” OR “Bolivia” OR “Cabo Verde” OR “Cambodia” OR “Cameroon” OR “Comoros” OR “Cote d’Ivoire” OR “Djibouti” OR “Egypt” OR “El Salvador” OR “Ghana” OR “Honduras” OR “India” OR “Papua New Guinea” OR “Indonesia” OR “Kenya” OR “Micronesia” OR “Kyrgyzstan” OR “Laos” OR “Lesotho” OR “Mauritania” OR “Moldova” OR “Mongolia” OR “Morocco” OR “Myanmar” OR “Nicaragua” OR “Nigeria” OR “Pakistan” OR “Philippines” OR “Sao Tome and Principe” OR “Senegal” OR “Melanesia” OR “Sudan” OR “Eswatini” OR “Timor-Leste” OR “Tunisia” OR “Ukraine” OR “Uzbekistan” OR “Vanuatu” OR “Vietnam” OR “Zambia” OR “Zimbabwe” OR “Albania” OR “Algeria” OR “American Samoa” OR “Argentina” OR “Armenia” OR “Azerbaijan” OR “Republic of Belarus” OR “Belize” OR “Bosnia and Herzegovina” OR “Botswana” OR “Brazil” OR “Bulgaria” OR “China” OR “Colombia” OR “Costa Rica” OR “Cuba” OR “Dominica” OR “Dominican Republic” OR “Equatorial Guinea” OR “Ecuador” OR “Fiji” OR “Gabon” OR “Georgia (Republic)” OR “Grenada” OR “Guatemala” OR “Guyana” OR “Iran” OR “Iraq” OR “Jamaica” OR “Jordan” OR “Kazakhstan” OR “Kosovo” OR “Lebanon” OR “Libya” OR “Republic of North Macedonia” OR “Malaysia” OR “Indian Ocean Islands” OR “Mexico” OR “Montenegro” OR “Namibia” OR “Paraguay” OR “Peru” OR “Russia” OR “Samoa” OR “Serbia” OR “Sri Lanka” OR “South Africa” OR “Saint Lucia” OR “Saint Vincent and the Grenadines” OR “Suriname” OR “Thailand” OR “Tonga” OR “Turkey” OR “Turkmenistan” OR “Venezuela” OR “Mozambique” OR “Eritrea” OR “Guinea-Bissau” OR “Congo”))) **AND** (“Randomized Controlled Trial” [Publication Type] OR “Clinical Trial” [Publication Type] OR “Controlled Clinical Trial” [Publication Type] OR “randomized controlled trial” OR rct OR “controlled trial”) AND (english[Filter] OR french[Filter] OR portuguese[Filter] OR spanish[Filter])

#### B. Physiotherapy Evidence Database (PEDro)

Abstract &Title: robot* stroke*

Subdiscipline: neurology

Method: clinical trial

When searching: Match all search terms (AND)

Number of hits: 165

Abstract &Title: tele* stroke*

Subdiscipline: neurology

Method: clinical trial

When searching: Match all search terms (AND)

Number of hits: 47

Abstract &Title: sensor* stroke*

Subdiscipline: neurology

Method: clinical trial

When searching: Match all search terms (AND)

Number of hits: 166

Abstract &Title: game* stroke*

Subdiscipline: neurology Method: clinical trial

When searching: Match all search terms (AND)

Number of hits: 58

Abstract &Title: virtual reality *stroke

Subdiscipline: neurology

Method: clinical trial

When searching: Match all search terms (AND)

Number of hits: 99

Total search: 535

A hand search will be performed against the previous search strategy to select studies that include upper limb assessment and the low- and middle-income countries described in the PubMed search.

#### C. Global Index Medicus

##### Descriptor 1: Stroke

Results: 43 657

tw:(tw:((tw:(stroke)) OR (tw:(stroke*)) OR (tw:(cerebrovascular accident*)) OR (tw:(cerebrovascular accidents)) OR (tw:(cva)) OR (tw:(cvas)) OR (tw:(cerebrovascular apoplexy)) OR (tw:(cerebrovascular* apoplex*)) OR (tw:(brain vascular accident)) OR (tw:(brain vascular accidents)) OR (tw:(brain* vascular* accident*)) OR (tw:(cerebrovascular stroke)) OR (tw:(cerebrovascular* stroke*)) OR (tw:(cerebrovascular strokes)) OR (tw:(apoplexy)) OR (tw:(aplopex*)) OR (tw:(cerebral stroke)) OR (tw:(cerebral strokes)) OR (tw:(acute stroke)) OR (tw:(acute strokes)) OR (tw:(chronic stroke)) OR (tw:(chronic strokes)) OR (tw:(subacute stroke)) OR (tw:(subacute strokes)) OR (tw:(acute cerebrovascular accident)) OR (tw:(acute* cerebrovascular* accident*)) OR (tw:(paresis)) OR (tw:(hemiplegia)) OR (tw:(brain infarction)) OR (tw:(cerebrovascular disorder)) OR (tw:(cerebral infarction)) OR (tw:(brain ischemia)) OR (tw:(cerebral ischemia))))

##### Descriptor 2: Rehabilitation

Results: 68 500

tw:((tw:(rehabilitation)) OR (tw:(“physical and rehabilitation medicine”)) OR (tw:(stroke rehabilitation)) OR (tw:(neurological rehabilitation)) OR (tw:(orthopedic procedure*)) OR (tw:(telerehabilitation)) OR (tw:(tele-rehabilitation*)) OR (tw:(remote rehabilitation)) OR (tw:(virtual rehabilitation)) OR (tw:(“activities of daily living”)) OR (tw:(exercise therapy)) OR (tw:(rehabilitation center*)) OR (tw:(recreation therapy)) OR (tw:(“recovery of function”)) OR (tw:(occupational therapy)) OR (tw:(neurorehabilitation)) OR (tw:(physiotherapy)))

##### Descriptor 3: Technology

Results: 46 340

tw:((tw:(virtual reality)) OR (tw:(virtual reality exposure)) OR (tw:(virtual reality exposure therapy)) OR (tw:(robotic)) OR (tw:(robotics)) OR (tw:(robot*)) OR (tw:(robots)) OR (tw:(video game)) OR (tw:(video games)) OR (tw:(telerehabilitation)) OR (tw:(technology)) OR (tw:(remote sensing technology)) OR (tw:(wireless technology)) OR (tw:(biomedical technology)) OR (tw:(electronics)) OR (tw:(mobile applications)) OR (tw:(remote consultation)) OR (tw:(telemedicine)) OR (tw:(robotics rehabilitation)) OR (tw:(robot rehabilitation)) OR (tw:(telerobotics)) OR (tw:(robot-assisted rehabilitation)) OR (tw:(robot device)) OR (tw:(computer game)) OR (tw:(computer games)) OR (tw:(electronic device*)) OR (tw:(electronic devices)) OR (tw:(e-device)))

###### Final Search (Descriptor 1 + 2 + 3)

Results: 230

tw:((tw:(tw:(tw:((tw:(stroke)) OR (tw:(stroke*)) OR (tw:(cerebrovascular accident*)) OR (tw:(cerebrovascular accidents)) OR (tw:(cva)) OR (tw:(cvas)) OR (tw:(cerebrovascular apoplexy)) OR (tw:(cerebrovascular* apoplex*)) OR (tw:(brain vascular accident)) OR (tw:(brain vascular accidents)) OR (tw:(brain* vascular* accident*)) OR (tw:(cerebrovascular stroke)) OR (tw:(cerebrovascular* stroke*)) OR (tw:(cerebrovascular strokes)) OR (tw:(apoplexy)) OR (tw:(apoplex*)) OR (tw:(cerebral stroke)) OR (tw:(cerebral strokes)) OR (tw:(acute stroke)) OR (tw:(acute strokes)) OR (tw:(chronic stroke)) OR (tw:(chronic strokes)) OR (tw:(subacute stroke)) OR (tw:(subacute strokes)) OR (tw:(acute cerebrovascular accident)) OR (tw:(acute* cerebrovascular* accident*)) OR (tw:(paresis)) OR (tw:(hemiplegia)) OR (tw:(brain infarction)) OR (tw:(cerebrovascular disorder)) OR (tw:(cerebral infarction)) OR (tw:(brain ischemia)) OR (tw:(cerebral ischemia)))))) AND (tw:(tw:((tw:(rehabilitation)) OR (tw:(“physical and rehabilitation medicine”)) OR (tw:(stroke rehabilitation)) OR (tw:(neurological rehabilitation)) OR (tw:(orthopedic procedure*)) OR (tw:(telerehabilitation)) OR (tw:(tele-rehabilitation*)) OR (tw:(remote rehabilitation)) OR (tw:(virtual rehabilitation)) OR (tw:(“activities of daily living”)) OR (tw:(exercise therapy)) OR (tw:(rehabilitation center*)) OR (tw:(recreation therapy)) OR (tw:(“recovery of function”)) OR (tw:(occupational therapy)) OR (tw:(neurorehabilitation)) OR (tw:(physiotherapy))))) AND (tw:(tw:((tw:(virtual reality)) OR (tw:(virtual reality exposure)) OR (tw:(virtual reality exposure therapy)) OR (tw:(robotic)) OR (tw:(robotics)) OR (tw:(robot*)) OR (tw:(robots)) OR (tw:(video game)) OR (tw:(video games)) OR (tw:(telerehabilitation)) OR (tw:(technology)) OR (tw:(remote sensing technology)) OR (tw:(wireless technology)) OR (tw:(biomedical technology)) OR (tw:(electronics)) OR (tw:(mobile applications)) OR (tw:(remote consultation)) OR (tw:(telemedicine)) OR (tw:(robotics rehabilitation)) OR (tw:(robot rehabilitation)) OR (tw:(telerobotics)) OR (tw:(robot-assisted rehabilitation)) OR (tw:(robot device)) OR (tw:(computer game)) OR (tw:(computer games)) OR (tw:(electronic device*)) OR (tw:(electronic devices)) OR (tw:(e-device))))))

